# Independently ambulatory children with spina bifida experience near-typical joint moments and forces during walking

**DOI:** 10.1101/2022.06.02.22275843

**Authors:** Marissa R. Lee, Jennifer L. Hicks, Tishya A. L. Wren, Scott L. Delp

## Abstract

**Background:** Spina bifida, a neurological defect, can result in lower-limb muscle weakness. Altered ambulation and reduced musculoskeletal loading can yield decreased bone strength in individuals with spina bifida, yet individuals who remain ambulatory can exhibit normal bone outcomes.

**Research question:** During walking, how do lower-limb joint kinematics, moments, and forces in independently ambulatory children with spina bifida differ from those of children with typical development?

**Methods:** We retrospectively analyzed data from 16 independently ambulatory children with spina bifida and 16 children with typical development and confirmed that bone strength was similar between the two groups. Plantar flexor muscle strength was measured by manual muscle testing, and 14 of the children with spina bifida wore activity monitors for one week. We estimated joint forces using motion capture data and musculoskeletal simulations. We used Statistical Parametric Mapping *t*-tests to compare lower-limb joint kinematic and kinetic waveforms between the groups with spina bifida and typical development. Within the group with spina bifida, we examined relationships between plantar flexor muscle strength and peak tibial forces by calculating Spearman correlations.

**Results:** Activity monitors from the children with spina bifida reported typical daily steps (9656 [SD 3095]). Despite slower walking speeds (p=0.004) and altered lower-body kinematics (p<0.0001), children with spina bifida had joint moments and forces similar to those of children with typical development, with no detectable differences during stance. Plantar flexor muscle weakness was associated with increased compressive knee force (p=0.001) and shear ankle force (p=0.006).

**Significance:** High-functioning, independently ambulatory children with spina bifida exhibited near-typical bone strength and near-typical step counts and load magnitudes. Our results suggest that maintaining ambulation and muscle strength can promote bone health in this population.

**Highlights:**

- Gait analysis was performed in high-functioning children with spina bifida
- On average, these children had typical knee & ankle moments & forces during walking
- Weak plantar flexor muscles were associated with increased tibial forces

## Introduction

Spina bifida is a neurological birth defect resulting from incomplete closure of the spinal column in early fetal development. In many cases, insufficient protection causes damage to the spinal cord and nerves. As a result, muscles in the lower extremities can be weakened or paralyzed. With increasing severity, spina bifida weakens the plantar flexor, gluteal, and quadriceps muscles [1]. In the US, approximately 166,000 individuals live with spina bifida [2]. Worldwide, it occurs in 2 to 8 of every 1000 live births [3].

Children with spina bifida have diminished motor abilities [4] and spend less time being active [5] than peers with typical development, in part due to lower-limb muscle weakness. These activity differences are hypothesized to contribute to the population’s increased risk of osteoporosis and low-energy fracture in lower-extremity weight-bearing long bones [6–9]. These increased risks are not constant across the population with spina bifida. With increasing ambulation and function, individuals with spina bifida have better bone health, including increased bone strength properties in the tibia [9,10] and femoral neck [11], increased tibia bone mass and volume [12], more normal tibial development [13], and lower risk for bone fracture [8]. In studying bone outcomes in individuals with spina bifida, it is important to understand why individuals with higher levels of function and ambulation demonstrate better bone health. This understanding will provide insight into how we might preserve bone health in this population.

Bone accrual and maintenance are influenced by mechanical loading. Dynamic loads drive bone adaptation, with loading magnitude generally regarded as more important than number of load cycles [14]. Insufficient mechanical stimulation of the bone yields bone resorption and lower bone strength [15]. Challenges in directly measuring joint forces have limited our ability to quantitatively understand bone loading. Fortunately, advances in musculoskeletal simulation enable joint force estimation [16–19].

In this study, we investigated joint loads during walking gait in high-functioning, independently ambulatory children with spina bifida. Walking is a daily activity that is impacted by spina bifida, and altered gait patterns affect joint loading. For example, lower-limb joint loads can be reduced with slower walking [18] or when smaller muscle forces are generated [19]. To investigate these issues, we sought to answer two questions: 1) How do lower-limb joint kinematics, moments, and forces during gait differ between independently ambulatory children with spina bifida and children with typical development? 2) Are there relationships between muscle strength and tibial forces in independently ambulatory children with spina bifida? By exploring these questions, we hope to understand bone loading during walking and how joint forces relate to muscle strength in high-functioning children with spina bifida. Our results provide insight into how daily mechanical loading relates to bone health in this population.

## Methods

### Data Collection

Data for this study were previously collected at Children’s Hospital Los Angeles [9,12]. This dataset contains demographic, step count, physical exam, computed tomography, and gait data of 83 ambulatory children with spina bifida and 179 children with typical development. The current analysis included only participants who could walk independently and barefoot, without orthoses or walking aids like crutches, and who had at least one complete gait cycle of kinematic and kinetic walking data (three consecutive force plate strikes, with a single foot contacting each plate).

Sixteen independent ambulators with spina bifida (mean age 10.4, standard deviation [SD] 2.7 years old, 8 female / 8 male) and 16 age- and sex-matched individuals with typical development were included (Table 1). The group with spina bifida comprised 11 individuals with myelomeningocele and 5 with lipomyelomeningocele. Using the Dias functional classification of myelomeningocele [1], physical therapists classified 2 individuals with spina bifida with high sacral functional levels and 14 with low sacral functional levels. For each included individual with spina bifida, an individual with typical development was matched such that the sexes were the same and the ages were as close as available. No significant differences (p≥0.868) in age, height, weight, or body mass index (BMI) existed between the groups with spina bifida and typical development. Written informed assent and consent were obtained from all participants and their guardians. All study procedures were approved by our institutional review boards.

**Table 1.**
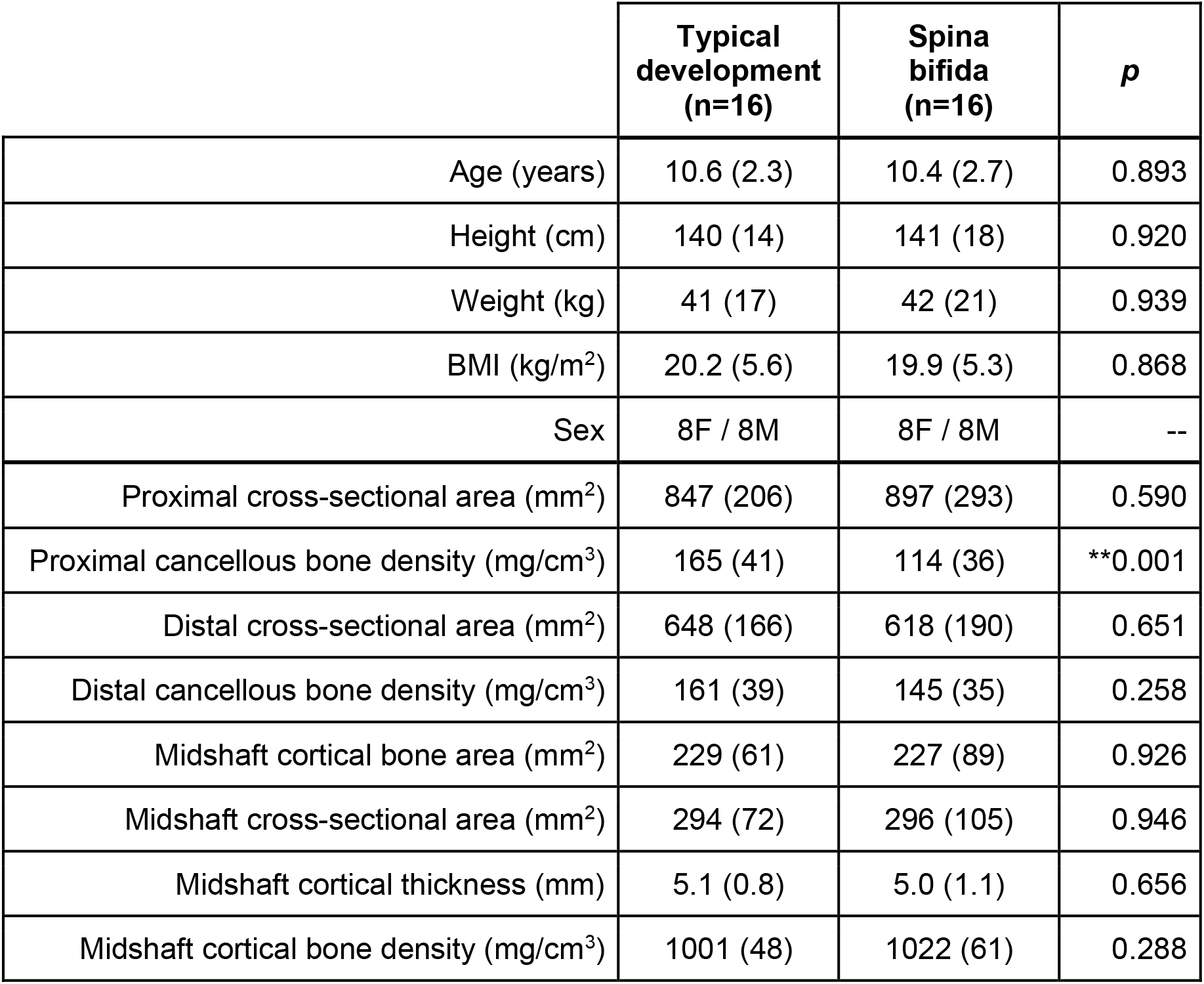
Group characteristics. Mean (standard deviation) characteristics within the groups with typical development and spina bifida and *p*-values of a Welch’s *t*-test between groups. (BMI=body mass index; **p<0.01)

Bone strength outcomes for all participants were calculated from quantitative computed tomography scans of each participant’s tibias. Image acquisition and processing procedures are described in a previous study [9]. Of eight bone properties measured, only cancellous bone density at the proximal tibia (p=0.001) was significantly lower (p<0.05) in the group with spina bifida than in the group with typical development (Table 1).

Individuals with spina bifida underwent exams with a physical therapist. Plantar flexor muscle strength was evaluated using a standard Manual Muscle Test (MMT, 0-5 scale [20]). Among participants with spina bifida, 14 of 16 wore StepWatch activity monitors (Modus, Edmonds, WA) for 1-9 (mean 5, median 6) days. Average steps per day were calculated for each participant.

Overground gait data of participants walking barefoot and without assistance at self-selected speeds were obtained for 1 to 5 gait cycles per participant (mean 2.1, median 2). Data from every complete gait cycle of kinematic and kinetic walking data (three consecutive force plate strikes, with a single foot contacting each plate) were included. Ground reaction forces were acquired at 2520 Hz through three consecutive floor-embedded force plates (AMTI, Watertown, MA) and were filtered at 6 Hz. Marker data of the torso and lower extremities were acquired at 120 Hz by an 8-camera motion capture system (Vicon, Oxford, UK), following the Plug-in-Gait model, with thigh markers replaced by patella markers [21].

### Musculoskeletal Simulation

We estimated joint kinematics, moments, and forces using OpenSim 4.1 [22,23]. We first scaled a generic musculoskeletal model to match the size of each participant. The generic model was based on the full-body model developed by Rajagopal et al. [24] and modified by Uhlrich et al. [25]. Arms in the model were fixed to the torso, since arm marker data were not collected. We scaled peak isometric muscle forces to each participant using the mass-height-volume relationship estimated by Handsfield et al. [24,26]. Using OpenSim’s Inverse Kinematics tool, we calculated joint kinematics. Across gait cycles, average marker root mean square (RMS) error was 1.1 (SD 0.2) cm, and maximum marker error was 3.5 (SD 0.4) cm, within recommended accuracy [27]. We filtered kinematics at 6 Hz.

To reduce dynamic inconsistencies in the experimental data, we used the Residual Reduction Algorithm [22]. RMS and peak residual forces were 1.6% (SD 0.6%) and 3.5% (SD 1.7%) of the maximum ground reaction force magnitude, respectively, within the recommended limit of 5% (Hicks 2015). RMS and peak residual moments were 1.7% (SD 0.4%) and 3.7% (SD 1.2%) of the center of mass height times maximum ground reaction force magnitude, respectively. These moment discrepancies are greater than the 1% recommended discrepancy, likely because the arms were fixed to the torsos of our models [27,28].

We estimated muscle forces using a static optimization algorithm that minimized the sum of squared muscle activations. The algorithm accounted for passive muscle forces and tendon compliance [25]. Using the inverse dynamics and static optimization results, we summed intersegmental resultant forces and muscle forces to estimate shear and compressive forces on the tibia at the knee and ankle joints.

### Musculoskeletal Simulation Evaluation

To evaluate the ability of our simulations to represent muscle activity, we qualitatively compared simulation muscle activation estimates to electromyography (EMG) recordings in a separate group of independently ambulatory participants. We performed this comparison with a separate group, comprising 1 participant with typical development and 6 participants with spina bifida, because EMG data were not collected in the primary cohort. Overground walking EMG recordings from the gastrocnemius medialis, tibialis anterior, semitendinosus, rectus femoris, and vastus lateralis were acquired at 2400 Hz, band-pass filtered between 50 and 500 Hz, rectified, low-pass filtered at 7.5 Hz, and normalized such that peak EMG signals matched peak simulated activations.

We qualitatively compared muscle activations computed using static optimization and EMG signals to determine if major features of experimental EMG were represented in our simulations (Supplementary Figure 1). Both simulated muscle activity and EMG recordings indicated gastrocnemius activity in late stance; tibialis anterior activity in early stance and swing; semitendinosus activity in early stance and late swing; and vastus lateralis activity in early stance. Peak static optimization activation estimates of the gastrocnemius medialis tended to decrease with plantar flexor muscle strength.

### Statistical Analysis

We compared walking speed between the groups with spina bifida and typical development using Welch’s *t*-test.

We normalized internal joint moments by body mass and normalized joint forces by body weight. For each participant with spina bifida, we analyzed only the leg with weaker plantar flexor muscle strength. If both legs were equally affected, and for participants with typical development, we analyzed the leg with more available gait cycles. For each selected limb, we extracted and averaged gait waveforms (lower-limb joint kinematics and moments, ground reaction forces, and tibial forces at the knee and ankle) across gait cycles.

To determine how lower-limb gait waveforms differed between independently ambulatory children with spina bifida and children with typical development, we compared average gait waveforms using Statistical Parametric Mapping (SPM) unpaired *t*-tests. This technique [29] enables the comparison of the means of two groups of continuous waveforms and identification of periods of difference in the gait cycle. It has previously been used to identify differences in walking gait patterns in children with cerebral palsy [30]. We also used Welch’s *t*-tests to compare kinetic peaks between groups.

To determine if relationships exist between muscle function and tibial forces, we calculated the Spearman correlation between plantar flexor muscle strength and peak tibial forces. Manual Muscle Test scores that involved + or - were replaced with ±0.33 adjustments (e.g., 3+ re-coded as 3.33, 3-re-coded as 2.67).

All statistical tests were evaluated with a significance level of 0.05. Statistical analyses were conducted in Python 3 (Natick, MA, USA).

Data and custom simulation and analysis scripts are available on SimTK at: https://simtk.org/projects/sb_force.

## Results

### Walking Activity and Speed

Activity monitors worn by 14 participants with spina bifida reported 9656 (SD 3095) average daily steps. Average walking speed, measured from the gait analysis trials, was slower (p=0.004) in the group with spina bifida (1.1 [SD 0.1] m/s) than in the group with typical development (1.3 [SD 0.1] m/s).

### Gait Waveforms

The group with spina bifida exhibited significantly greater ankle dorsiflexion angles in the second half of stance and lower plantarflexion angles at toe-off (p<0.0001) and significantly lower knee flexion angles at toe-off (p=0.032) compared to the group with typical development. Hip flexion and adduction angles did not differ significantly between groups. Greater variances in joint kinematics among participants with spina bifida were observed across the gait cycle at the hip, knee, and ankle (Figure 1).

**Figure 1.**
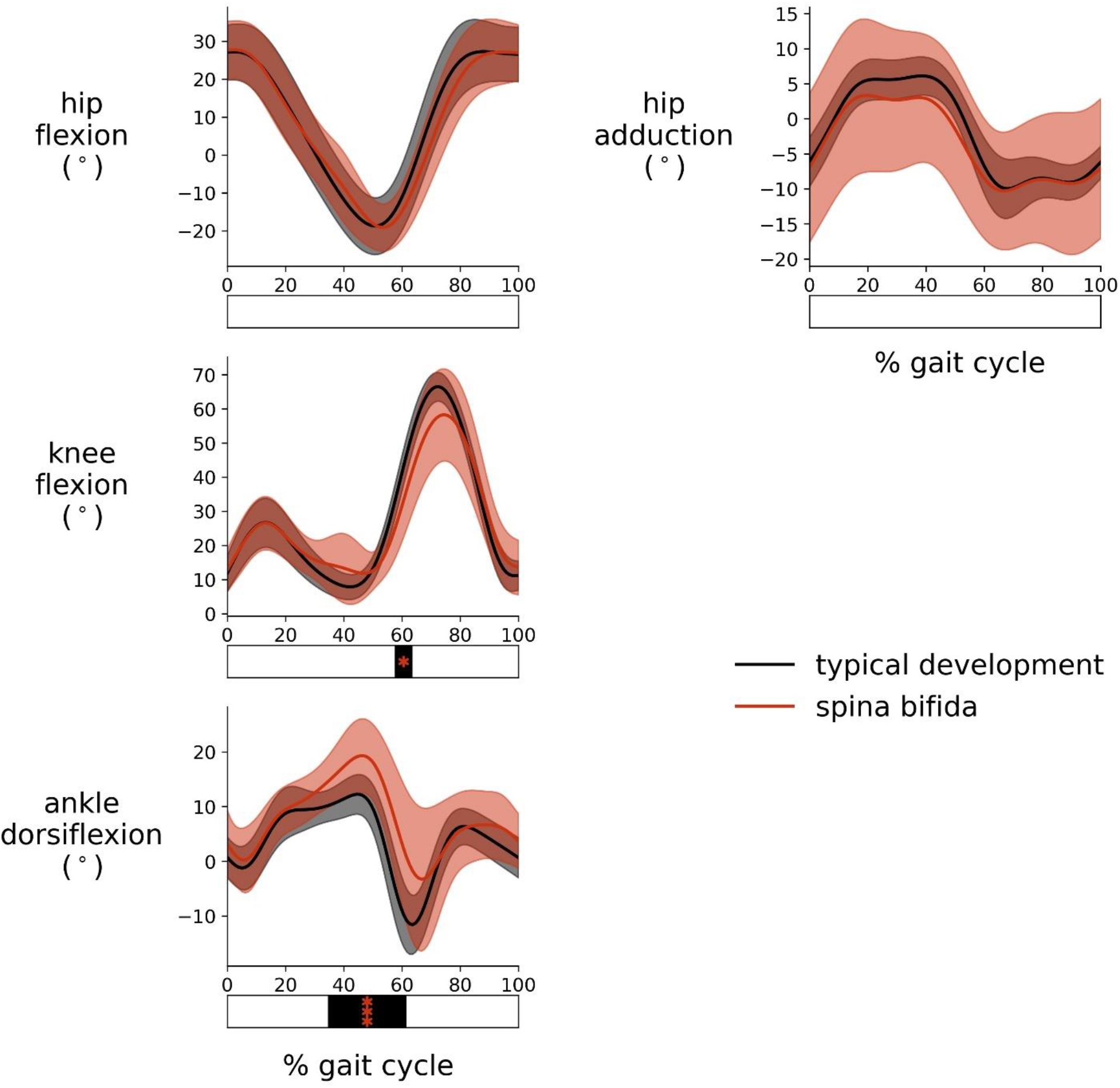
Group joint kinematic waveforms. Mean ± one standard deviation joint kinematics across the gait cycle for participants in the groups with typical development (black) and spina bifida (red). Bars underneath the curves reflect significant difference results from the SPM *t*-tests (*p<0.05, ***p<0.001).

Few statistically significant differences in joint moments were identified between groups (Figure 2). Differences in hip extension moments (p=0.025), hip abduction moments (p=0.040), and knee extension moments (p=0.037) occurred only in swing and for periods less than 4% of the gait cycle. As with the kinematics, joint moment variances were greater in the group with spina bifida than in the group with typical development. Mean ankle plantarflexion moment during stance trended lower in the group with spina bifida, but no significant differences were detected. Additionally, no significant differences were detected in the peak values of these joint moment waveforms (p>0.098).

**Figure 2.**
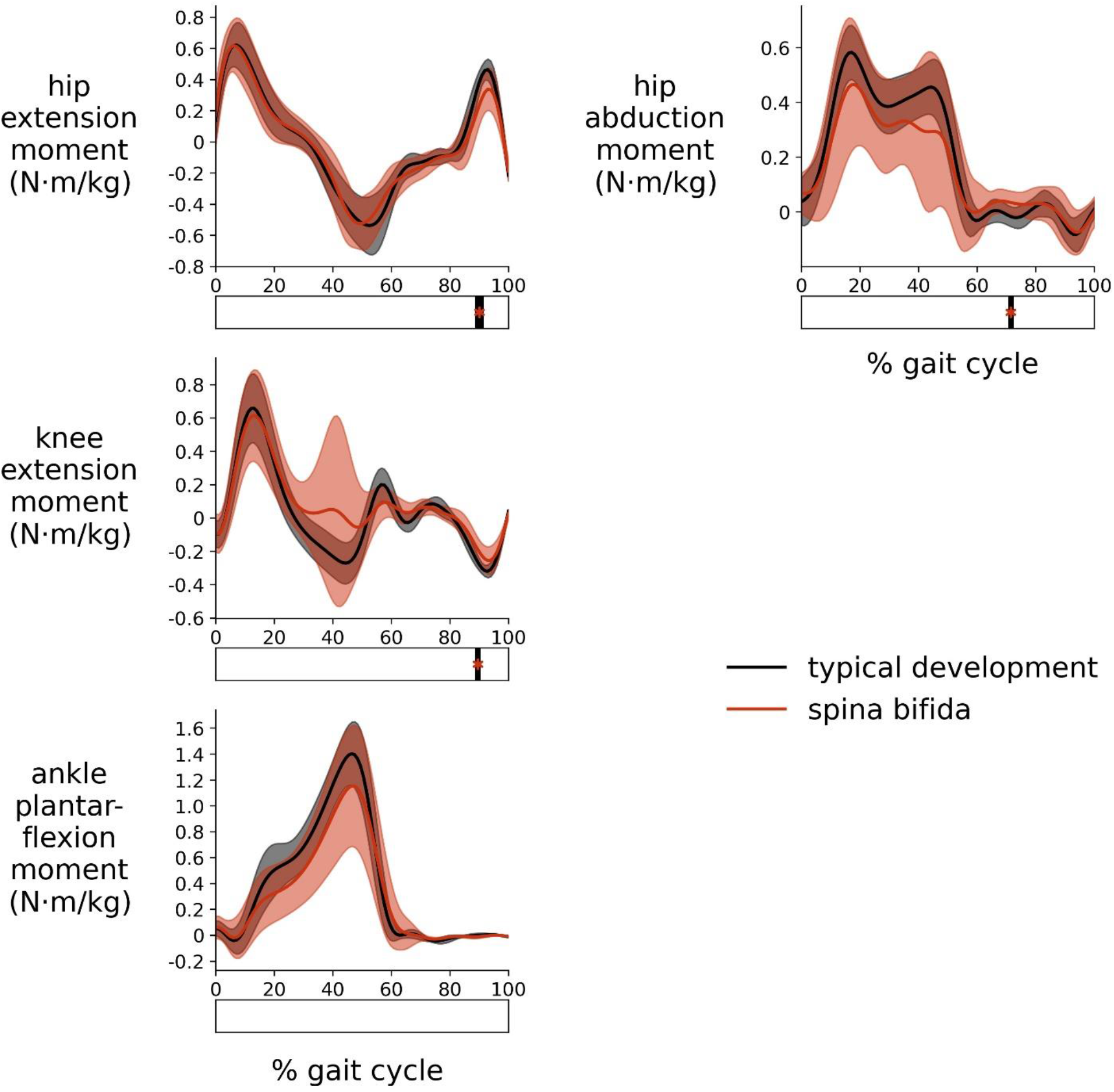
Group joint moment waveforms. Mean ± one standard deviation joint moments across the gait cycle for participants in the groups with typical development (black) and spina bifida (red). Joint moments are normalized by body mass. Bars underneath the curves reflect significant difference results from the SPM *t*-tests (*p<0.05).

No differences between the two groups were detected in mean ground reaction forces (Figure 3) or in shear, compressive, or total tibial forces at the knee and ankle (Figure 4). Additionally, no significant differences were detected in the peak values of these joint force waveforms (p>0.133).

**Figure 3.**
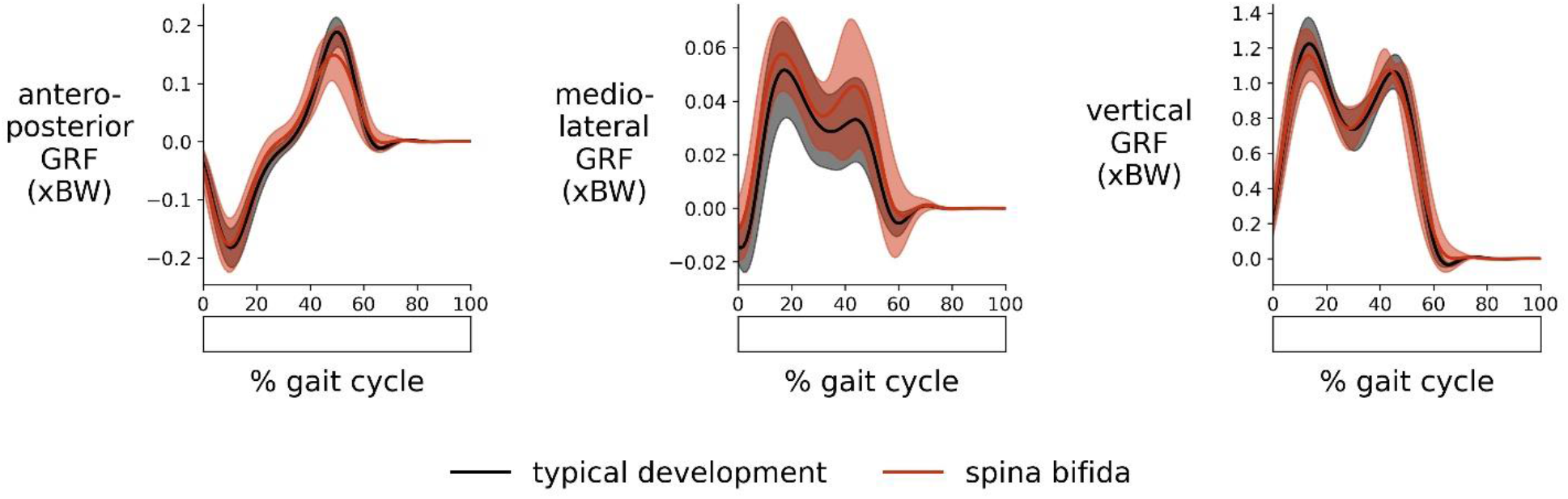
Group ground reaction force waveforms. Mean ± one standard deviation ground reaction forces (GRFs) across the gait cycle for participants in the groups with typical development (black) and spina bifida (red). GRFs are expressed in terms of number of body weights (xBW). No significant differences were detected between groups.

**Figure 4.**
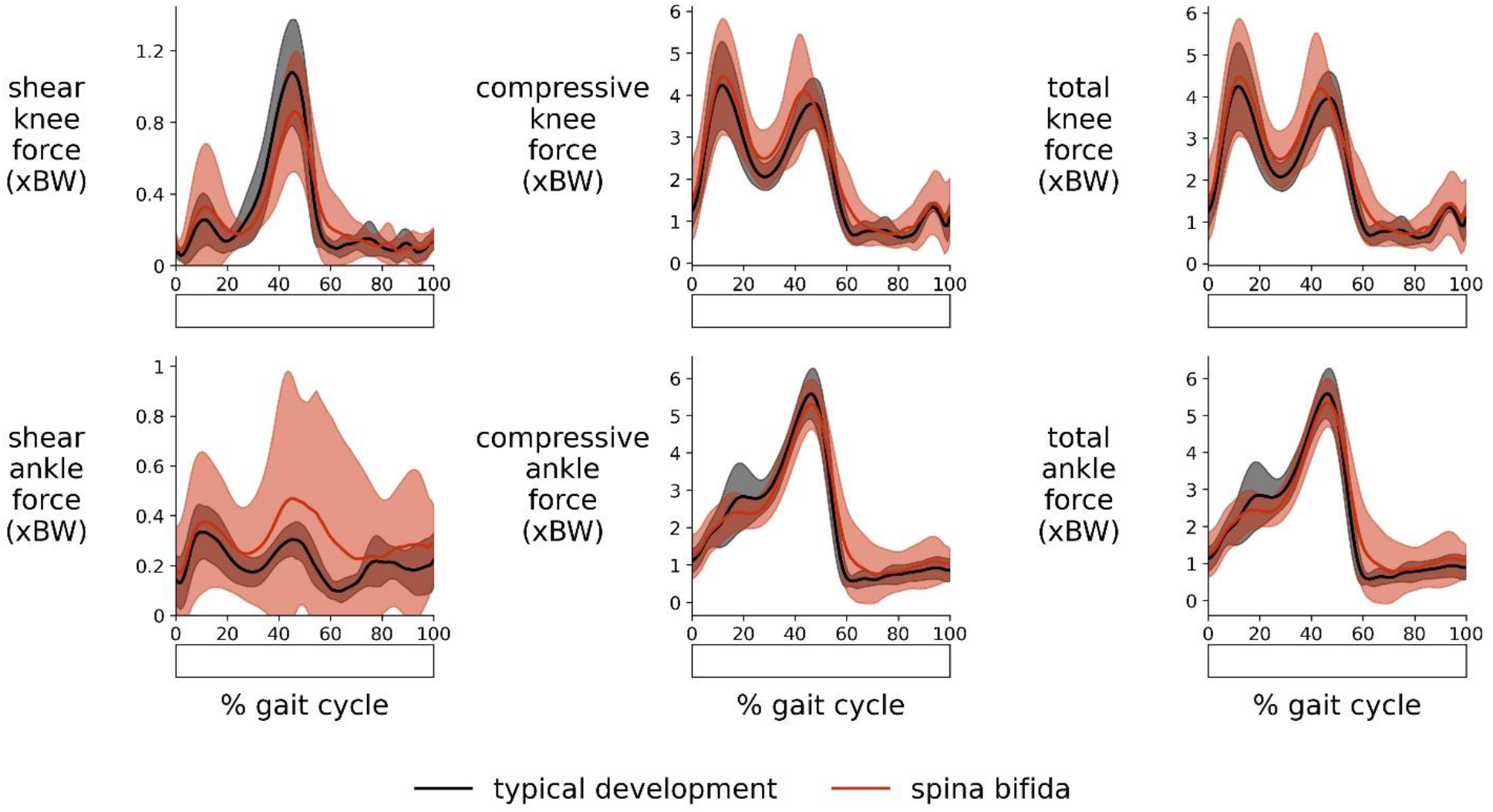
Group joint force waveforms. Mean ± one standard deviation joint forces across the gait cycle for participants in the groups with typical development (black) and spina bifida (red). Forces are expressed in terms of number of body weights (xBW). No significant differences were detected between groups.

### Plantar Flexor Muscle Strength and Tibial Forces

Plantar flexor muscle strength was negatively correlated with compressive knee force (*ρ* =-0.73, p=0.001) and shear ankle force (*ρ* =-0.66, p=0.006) in the group with spina bifida (Figure 5). The three individuals with the lowest plantar flexor muscle strength exhibited peak compressive knee forces and peak shear ankle forces greater than those exhibited by participants with typical development (mean + two standard deviations).

**Figure 5.**
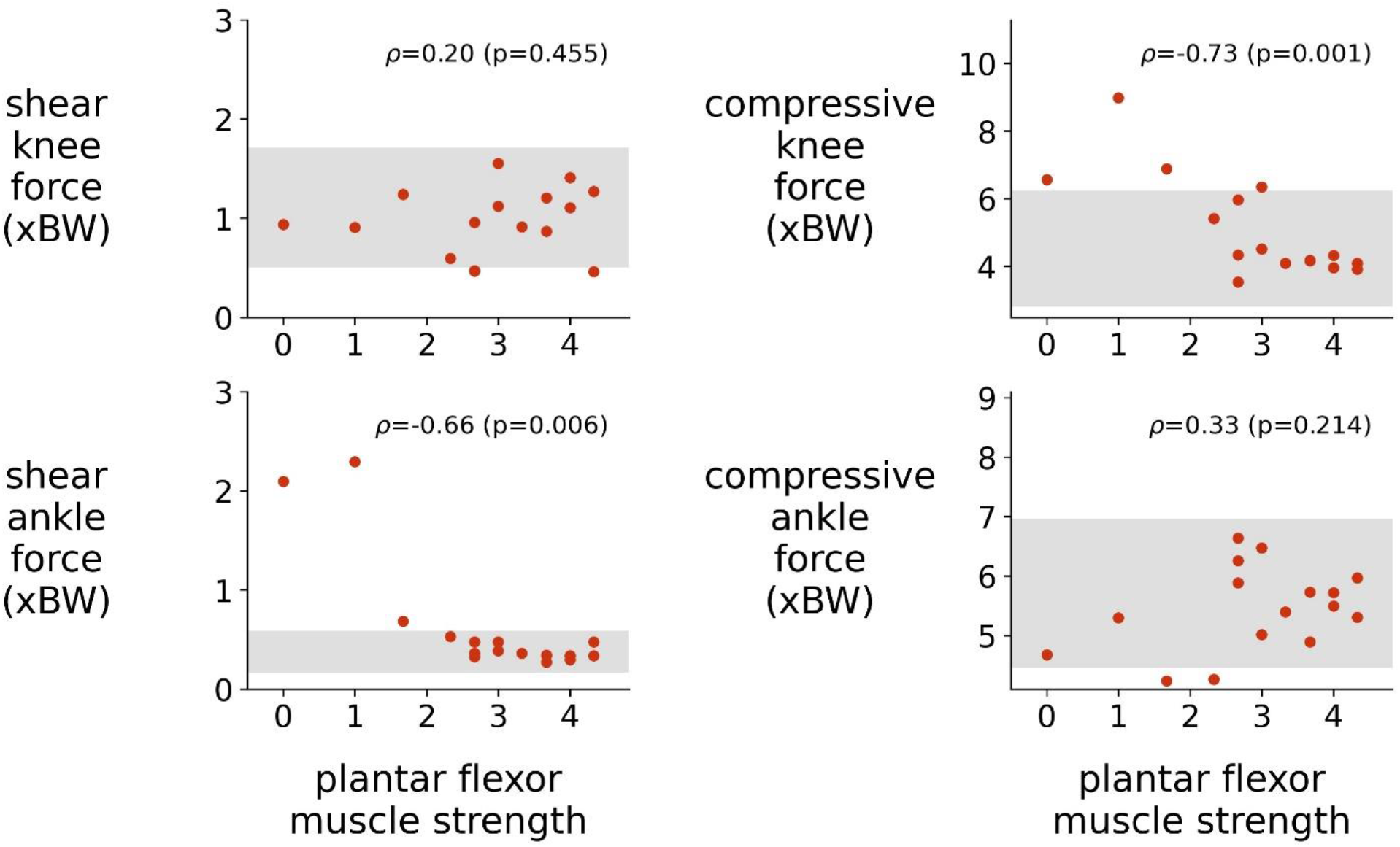
Peak joint force and plantar flexor muscle strength correlations. Forces are expressed in terms of number of body weights (xBW). Each data point represents the analyzed limb of a single participant. Each gray band represents the peak joint force mean ± two standard deviations of participants in the group with typical development. Spearman’s *ρ* was calculated for each relationship.

## Discussion

On average, children with spina bifida with near-typical bone health demonstrated near-typical lower-limb joint moments and forces, despite having slower walking speeds and altered lower-limb kinematics. We observed greater kinematic and kinetic variability across the gait cycle within the group with spina bifida. Variability in peak shear and compressive joint forces could be explained in part by differences associated with plantar flexor muscle weakness. Our results suggest that maintaining ambulation and muscle strength can promote bone health in children with spina bifida.

### Joint Kinematics & Moments

Previous studies have presented qualitative comparisons of joint kinematic and moment waveforms between groups with spina bifida and typical development [31,32]. In this study, we additionally analyzed continuous differences across the gait cycle using SPM *t*-tests. These tests enable identification of statistically significant periods of difference and account for variance in waveforms.

Independently ambulatory participants with spina bifida demonstrated near-typical kinematics on average. The main difference between groups occurred in ankle angles. Our findings of greater ankle dorsiflexion angles and lower plantarflexion angles are consistent with the ankle plantar flexor muscle weakness associated with spina bifida and with previous findings [31].

Despite differences in lower-limb kinematics, the group with spina bifida demonstrated near-typical joint moments. Gutierrez et al. [32] qualitatively compared joint moments among groups with typical development and varying levels of spina bifida. They reported decreased hip abduction and ankle plantarflexion moments during stance and decreased knee flexion moment in late stance. These differences, consistent with the weak hip abductor and plantar flexor muscles present in children with spina bifida, are supported by trends in our data, but we did not detect statistically significant differences in our high-functioning study group.

Average walking speed was slower than typical in the group with spina bifida. Schwartz et al. [33] reported joint kinematic and moment changes associated with walking speed in children with typical development. The directions of the trends we observed in our analyses align with those associated with decreased walking speed. Notably, however, the differences we observed between groups were greater than the reported changes due to slower walking speed alone.

### Tibial Forces & Bone Health

On average, our simulations estimated typical shear and compressive tibial forces in the group with spina bifida. This group also exhibited typical numbers of daily loading cycles (steps per day). The daily steps recorded on activity monitors worn by participants with spina bifida (9656 [SD 3095] steps) were comparable to those previously reported from 7 children with typical development who underwent the same activity monitoring protocol (9589 [SD 3322] steps [34]). Our findings of near-typical joint forces and loading cycles, combined with the near-typical bone health observed (Table 1), suggest that maintaining ambulation and muscle strength can promote bone health in this group. In general, this independently ambulatory population’s low risk for osteoporosis and fracture may result from near-typical mechanical loading patterns.

While we did not detect differences in the group means of the tibial forces, we did observe larger variation within the group with spina bifida. Variabilities may be partially attributable to variations in the ground reaction forces and joint configurations. They also likely reflect differences in muscle forces over the gait cycle, since muscles are the major contributing forces to joint forces [35]. Given this high variability, while on average independently ambulatory children with spina bifida do not have low loading or decreased bone strength, altered loading and low bone mass may still be present for some individuals with spina bifida.

Our investigations of the relationships between muscle and joint forces identified that weaker plantar flexor muscles were significantly correlated with greater peak shear ankle and compressive knee forces. Of particular note are the increased forces experienced by the weakest participants with spina bifida. These increased forces could impact fracture risk and bone strength. For example, elevated shear ankle and compressive knee forces could lead to fracture [36] and osteoarthritis [37,38]. While we found that plantar flexor muscle strength was associated with joint forces, we did not detect (p>0.5) significant correlations between plantar flexor muscle strength and bone strength measures.

Our study included only highly functional, independently ambulatory children with spina bifida and is limited by its sample size. Given the small sample, that we did not detect significant differences in many of our tests does not indicate that we would not detect differences in a study with a larger sample size. Future research should conduct joint force analyses with a larger and more diverse sample of individuals with spina bifida. Previous work has demonstrated that bone health disparities between individuals with spina bifida and individuals with typical development increase with puberty [9] and disparities in function [10–13]. Including older independent ambulators with spina bifida, as well as a group of individuals with spina bifida with a greater range of functional abilities, will be critical to understanding how bone loading differs among individuals with spina bifida and relates to bone health. Future studies should also investigate joint forces beyond walking gait and in other free-living activities.

We conclude that on average, young, high-functioning individuals with spina bifida undergo near-typical lower-limb joint moments and forces. These loads support the near-typical bone health observed in this group. Special attention should be paid to high-functioning individuals with spina bifida who have weak plantar flexor muscles, as they may undergo increased joint forces. Our results suggest that maintaining ambulation and muscle strength can help to promote bone health in children with spina bifida.

## Data Availability

All data produced are available online at

https://simtk.org/projects/sb_force

## Acknowledgements

Funding: This work was supported by the National Institutes of Health [grant numbers 5R01HD059826, P41EB027060]; the Inventec Stanford Graduate Fellowship, Stanford, CA; and the Catalyst for Collaborative Solutions, Stanford, CA. The study sponsors had no involvement in the study design; in the collection, analysis, or interpretation of data; in the writing of the manuscript; or in the decision to submit the manuscript for publication.

Our thanks to the team members of the Children’s Hospital Los Angeles John C. Wilson, Jr. Motion and Sports Analysis Laboratory for their collection and sharing of the data and to Scott Uhlrich and Carmichael Ong for their simulation expertise.

---

**Supplementary Figure 1.**
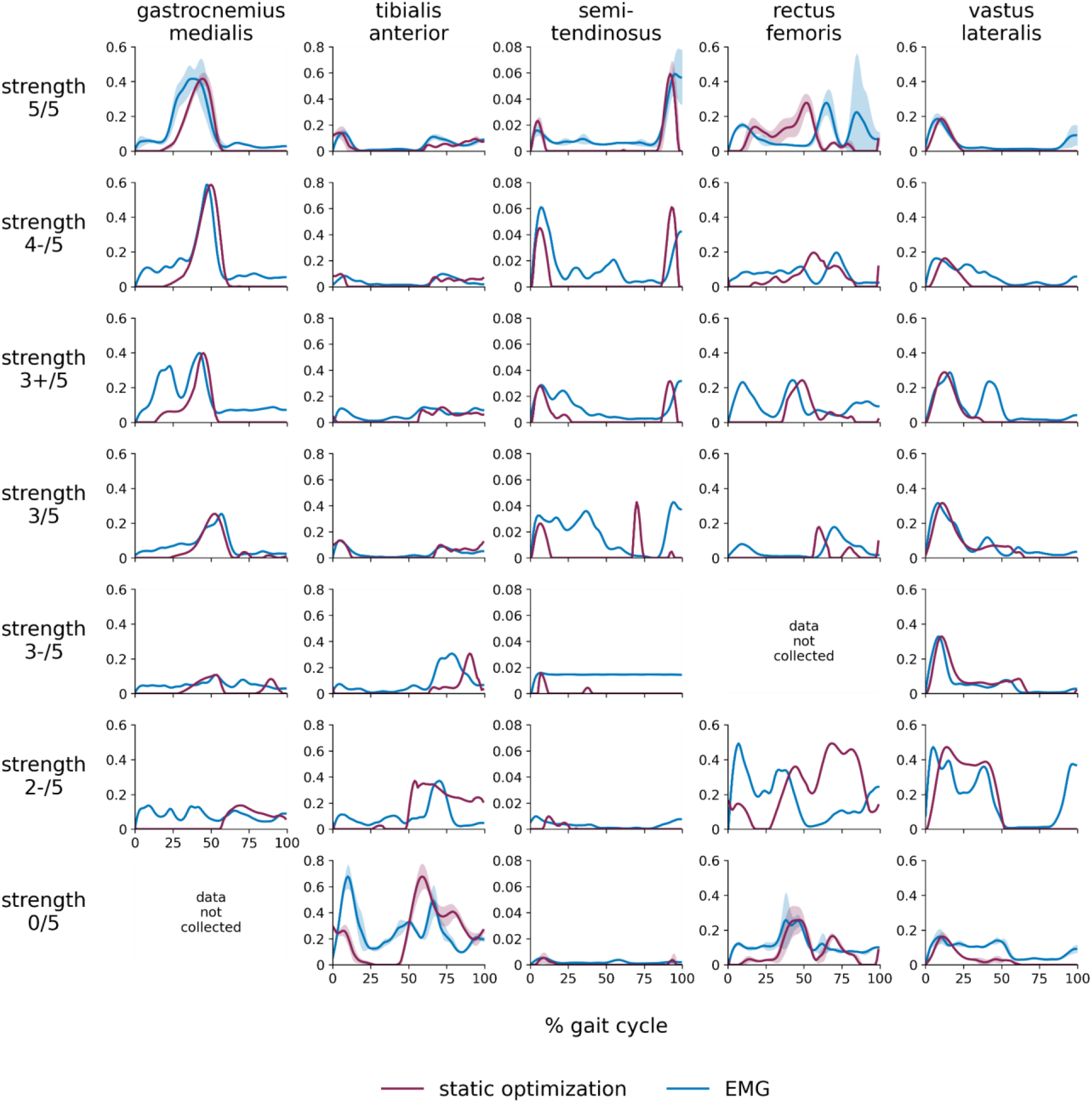
Comparison of muscle activations from static optimization to EMG signals. Mean ± one standard deviation muscle activations from static optimization (magenta) and filtered EMG recordings (blue) overlaid for five muscles of seven participants with varying plantar flexor muscle strengths. Participant results are ordered, from top to bottom, by plantar flexor muscle strength (the participant with typical development is on the top, followed by participants with spina bifida in order of decreasing plantar flexor muscle strength). The EMG recordings are normalized such that their peaks match those of the static optimization results.

## Ethics approval and consent to participate statement

This study was conducted with the approvals of the Children’s Hospital Los Angeles and Stanford Institutional Review Boards. Written informed assent and consent were obtained from all participants and their guardians.

## Data statement

The data and code supporting the conclusions of this study are available on SimTK at: https://simtk.org/projects/sb_force.

## Author statement

**Marissa R. Lee:** conceptualization, data curation, methodology, software, validation, formal analysis, investigation, visualization, writing - original draft. **Jennifer L. Hicks:** conceptualization, validation, writing - reviewing & editing, supervision, project administration, funding acquisition. **Tishya A. L. Wren:** conceptualization, data curation, methodology, resources, writing - reviewing & editing, supervision, funding acquisition. **Scott L. Delp:** conceptualization, writing - reviewing & editing, supervision, funding acquisition.

## References

[1] V.T. Swaroop, L. Dias, Orthopedic management of spina bifida. Part I: Hip, knee, and rotational deformities, J. Child. Orthop. 3 (2009) 441–449. https://doi.org/10.1007/s11832-009-0214-5.

[2] National Institute of Neurological Disorders and Stroke (NINDS), Spina bifida fact sheet, Bethesda, MD, 2013.

[3] A. Kondo, O. Kamihira, H. Ozawa, Neural tube defects: Prevalence, etiology and prevention, Int. J. Urol. 16 (2009) 49–57. https://doi.org/10.1111/j.1442-2042.2008.02163.x.

[4] M. Peny-Dahlstrand, A.-C. Åhlander, L. Krumlinde-Sundholm, G. Gosman-Hedström, Quality of performance of everyday activities in children with spina bifida: a population-based study, Acta Paediatr. 98 (2009) 1674–1679. https://doi.org/10.1111/j.1651-2227.2009.01410.x.

[5] E.A. Claridge, M.A.T. Bloemen, R.A. Rook, J. Obeid, B.W. Timmons, T. Takken, et al., Physical activity and sedentary behaviour in children with spina bifida, Dev. Med. Child Neurol. 61 (2019) 1400–1407. https://doi.org/10.1111/dmcn.14333.

[6] K. Parsch, Origin and treatment of fractures in spina bifida, Eur. J. Pediatr. Surg. 1 (1991) 298–306. https://doi.org/10.1055/s-2008-1042509.

[7] N.P. Dosa, M.F. Eckrich, D.A. Katz, M. Turk, G.S. Liptak, Incidence, prevalence, and characteristics of fractures in children, adolescents, and adults with spina bifida, J. Spinal Cord Med. 30 (2007) S5–S9. https://doi.org/10.1080/10790268.2007.11753961.

[8] M. Akbar, B. Bresch, P. Raiss, C.H. Fürstenberg, T. Bruckner, T. Seyler, et al., Fractures in myelomeningocele, J. Orthop. Traumatol. 11 (2010) 175–182. https://doi.org/10.1007/s10195-010-0102-2.

[9] T. al Wren, N.M. Mueske, S.A. Rethlefsen, R.M. Kay, A. van Speybroeck, W.J. Mack, Quantitative computed tomography assessment of bone deficits in ambulatory children and adolescents with spina bifida: Importance of puberty, JBMR Plus. 4 (2020). https://doi.org/10.1002/jbm4.10427.

[10] B.D. Rosenstein, W.B. Greene, R.T. Herrington, Bone density in myelomeningocele: The effects of ambulatory status and other factors, Dev. Med. Child Neurol. 29 (1987) 486– 494. https://doi.org/10.1111/j.1469-8749.1987.tb02508.x.

[11] S.D. Apkon, L. Fenton, J.R. Coll, Bone mineral density in children with myelomeningocele, Dev. Med. Child Neurol. 51 (2009) 63–67. https://doi.org/10.1111/j.1469-8749.2008.03102.x.

[12] R.E. Horenstein, S.J. Shefelbine, N.M. Mueske, C.L. Fisher, T.A.L. Wren, An approach for determining quantitative measures for bone volume and bone mass in the pediatric spina bifida population, Clin. Biomech. 30 (2015) 748–754. https://doi.org/10.1016/j.clinbiomech.2015.04.010.

[13] K.P. Chadwick, N.M. Mueske, R.E. Horenstein, S.J. Shefelbine, T.A.L. Wren, Children with myelomeningocele do not exhibit normal remodeling of tibia roundness with physical development, Bone. 114 (2018) 292–297. https://doi.org/10.1016/j.bone.2018.07.001.

[14] R.T. Whalen, D.R. Carter, C.R. Steele, Influence of physical activity on the regulation of bone density, J. Biomech. 21 (1988) 825–837. https://doi.org/10.1016/0021-9290(88)90015-2.

[15] D.R. Carter, G.S. Beaupré, Skeletal Function and Form, Cambridge University Press, Cambridge, 2000. https://doi.org/10.1017/CBO9780511574993.

[16] U. Glitsch, W. Baumann, The three-dimensional determination of internal loads in the lower extremity, J. Biomech. 30 (1997) 1123–1131. https://doi.org/10.1016/S0021-9290(97)00089-4.

[17] K.M. Steele, M.S. DeMers, M.H. Schwartz, S.L. Delp, Compressive tibiofemoral force during crouch gait, Gait Posture. 35 (2012) 556–560. https://doi.org/10.1016/j.gaitpost.2011.11.023.

[18] Z.F. Lerner, D.J. Haight, M.S. DeMers, W.J. Board, R.C. Browning, The effects of walking speed on tibiofemoral loading estimated via musculoskeletal modeling, J. Appl. Biomech. 30 (2014) 197–205. https://doi.org/10.1123/jab.2012-0206.

[19] M.S. DeMers, S. Pal, S.L. Delp, Changes in tibiofemoral forces due to variations in muscle activity during walking, J. Orthop. Res. 32 (2014) 769–776. https://doi.org/10.1002/jor.22601.

[20] M. Brown, D. Avers, Daniels and Worthingham’s Muscle Testing, tenth ed., Saunders, Philadelphia, 2018.

[21] T.A.L. Wren, K.P. Do, R. Hara, S.A. Rethlefsen, Use of a patella marker to improve tracking of dynamic hip rotation range of motion, Gait Posture. 27 (2008) 530–534. https://doi.org/10.1016/j.gaitpost.2007.07.006.

[22] S.L. Delp, F.C. Anderson, A.S. Arnold, P. Loan, A. Habib, C.T. John, et al., OpenSim: Open-source software to create and analyze dynamic simulations of movement, IEEE. Trans. Biomed. Eng. 54 (2007) 1940–1950. https://doi.org/10.1109/TBME.2007.901024.

[23] A. Seth, J.L. Hicks, T.K. Uchida, A. Habib, C.L. Dembia, J.J. Dunne, et al., OpenSim: Simulating musculoskeletal dynamics and neuromuscular control to study human and animal movement, PLoS Comput. Biol. 14 (2018) e1006223. https://doi.org/10.1371/journal.pcbi.1006223.

[24] A. Rajagopal, C.L. Dembia, M.S. DeMers, D.D. Delp, J.L. Hicks, S.L. Delp, Full-body musculoskeletal model for muscle-driven simulation of human gait, IEEE. Trans. Biomed. Eng. 63 (2016) 2068–2079. https://doi.org/10.1109/TBME.2016.2586891.

[25] S.D. Uhlrich, R.W. Jackson, A. Seth, J.A. Kolesar, S.L. Delp, Muscle coordination retraining inspired by musculoskeletal simulations reduces knee contact force, Sci. Rep. (2022, in press). https://doi.org/10.1038/s41598-022-13386-9.

[26] G.G. Handsfield, C.H. Meyer, J.M. Hart, M.F. Abel, S.S. Blemker, Relationships of 35 lower limb muscles to height and body mass quantified using MRI, J. Biomech. 47 (2014) 631–638. https://doi.org/10.1016/j.jbiomech.2013.12.002.

[27] J.L. Hicks, T.K. Uchida, A. Seth, A. Rajagopal, S.L. Delp, Is my model good enough? Best practices for verification and validation of musculoskeletal models and simulations of movement, J. Biomech. Eng. 137 (2015). https://doi.org/10.1115/1.4029304.

[28] M. Rosenberg, K.M. Steele, Simulated impacts of ankle foot orthoses on muscle demand and recruitment in typically-developing children and children with cerebral palsy and crouch gait, PLoS One. 12 (2017) e0180219. https://doi.org/10.1371/journal.pone.0180219.

[29] T.C. Pataky, Generalized n-dimensional biomechanical field analysis using statistical parametric mapping, J. Biomech. 43 (2010) 1976–1982. https://doi.org/10.1016/j.jbiomech.2010.03.008.

[30] A. Nieuwenhuys, E. Papageorgiou, K. Desloovere, G. Molenaers, T. de Laet, Statistical Parametric Mapping to identify differences between consensus-based joint patterns during gait in children with cerebral palsy, PLoS One. 12 (2017) e0169834. https://doi.org/10.1371/journal.pone.0169834.

[31] E.M. Gutierrez, Å. Bartonek, Y. Haglund-Åkerlind, H. Saraste, Characteristic gait kinematics in persons with lumbosacral myelomeningocele, Gait Posture. 18 (2003) 170–177. https://doi.org/10.1016/S0966-6362(03)00011-0.

[32] E.M. Gutierrez, Å. Bartonek, Y. Haglund-Åkerlind, H. Saraste, Kinetics of compensatory gait in persons with myelomeningocele, Gait Posture. 21 (2005) 12–23. https://doi.org/10.1016/j.gaitpost.2003.11.002.

[33] M.H. Schwartz, A. Rozumalski, J.P. Trost, The effect of walking speed on the gait of typically developing children, J. Biomech. 41 (2008) 1639–1650. https://doi.org/10.1016/j.jbiomech.2008.03.015.

[34] P. Yasmeh, N.M. Mueske, S. Yasmeh, D.D. Ryan, T.A.L. Wren, Walking activity during daily living in children with myelomeningocele, Disabil. Rehabil. 39 (2017) 1422–1427. https://doi.org/10.1080/09638288.2016.1198429.

[35] K.B. Shelburne, M.R. Torry, M.G. Pandy, Contributions of muscles, ligaments, and the ground-reaction force to tibiofemoral joint loading during normal gait, J. Orthop. Res. 24 (2006) 1983–1990. https://doi.org/10.1002/jor.20255.

[36] R. Flachsmann, N.D. Broom, A.E. Hardy, G. Moltschaniwskyj, Why is the adolescent moint particularly susceptible to osteochondral shear fracture?, Clin. Orthop. Relat. Res. 381 (2000) 212–221. https://doi.org/10.1097/00003086-200012000-00025.

[37] D.R. Carter, G.S. Beaupré, M. Wong, R.L. Smith, T.P. Andriacchi, D.J. Schurman, The mechanobiology of articular cartilage development and degeneration, Clin. Orthop. Relat. Res. 427 (2004) S69–S77. https://doi.org/10.1097/01.blo.0000144970.05107.7e.

[38] E.M. Roos, Joint injury causes knee osteoarthritis in young adults, Curr. Opin. Rheumatol. 17 (2005) 195–200. https://doi.org/10.1097/01.bor.0000151406.64393.00.

